# Why drinking episodes escalate differently: Event-level pathways linking hazardous alcohol consumption and sexual risk

**DOI:** 10.64898/2026.06.17.26355906

**Authors:** Thye Peng Ngo, Alexandrea Elizabeth Dunham, Glenn-Milo Santos

## Abstract

**Background:** Alcohol-involved drinking episodes vary in whether they involve hazardous alcohol consumption alone, near-miss sexual risk, or sexual risk behavior, but the within-event mechanisms underlying this variability remain unclear.

**Methods:** Guided by syndemic theory, we conducted a qualitative event-level analysis using modified grounded theory among adults in the San Francisco Bay Area who reported hazardous alcohol consumption, defined as an Alcohol Use Disorder Identification Test score ≥16. In-depth interviews elicited narratives of recent heavy drinking episodes and yielded 64 discrete drinking events across 22 participants. We focused on 35 events with evidence of within-event interaction between biopsychosocial and contextual factors. Using constant comparison, we identified escalation pathways, characterized interruption, and examined how events diverge into three outcomes: hazardous alcohol consumption only, hazardous alcohol consumption with near-miss sexual risk (when risk was plausible but not enacted), and hazardous alcohol consumption with sexual risk behavior.

**Results:** Two primary escalation pathways emerged. Dose-driven escalation involved cumulative alcohol or substance exposure that progressively impaired awareness and self-regulation. Meaning-driven escalation involved prioritizing connection, intimacy, or belonging despite awareness of risk. Time-driven continuation extended exposure across contexts and amplified both pathways. Hazardous alcohol consumption-only events more often followed dose-driven pathways, whereas events involving sexual risk behavior more often followed meaning-driven pathways. Near-miss events occurred across both pathways and illustrated how interruption before the escalation constraint point, when the capacity to modify behavior became reduced, could redirect escalation before sexual risk behavior occurred. Across events with similar levels of intoxication narratives, outcomes diverged according to when the interruption occurred and whether it altered escalation.

**Conclusion:** Hazardous drinking episodes diverge into different outcomes based on escalation pathways and the timing and effectiveness of interruption. Early and effective interruption before the escalation constraint point may represent a key target for harm-reduction strategies to prevent progression to sexual risk behavior.

## Introduction

Alcohol remains a major public health concern in the United States. In 2024, 50.6% of U.S. adults reported past-month alcohol use, and 21.7% reported past-month binge drinking, defined as consuming ≥5 drinks for men or ≥4 drinks for women on a single occasion; these estimates correspond to 132.6 million and 57.0 million adults, respectively [1–3]. Alcohol contributes to substantial morbidity and mortality, including physical and mental health consequences and harms to others [4–9]. Sexual behavior represents a particularly consequential domain of alcohol-related harm because risk often emerges within drinking episodes through moment-to-moment decisions rather than through stable individual risk alone [10,11].

A large body of research demonstrates associations between alcohol use and sexual risk behavior (SRB), including condomless sex, multiple sexual partners, and increased vulnerability to coercive or aggressive sexual encounters [12–15]. These risks are particularly important in episodes involving hazardous alcohol consumption (HAC), where heavier drinking may impair judgment, increase disinhibition, and reduce the feasibility of protective decision-making [16,17]. Experimental studies show that alcohol increases the likelihood of unprotected sex under conditions of sexual arousal [17]. Across adolescent and adult populations, systematic reviews and meta-analyses link alcohol use to condomless sex, earlier sexual initiation, and a greater number of sexual partners [12,18]. Among people living with HIV, alcohol use is associated with higher odds of condomless sex [19]. While these findings establish HAC and SRB as important domains of alcohol-related harm, most evidence is based on between-person or global associations [10,20], which obscure substantial variability in how alcohol-involved episodes unfold.

Event-level and daily diary studies provide a more nuanced perspective, showing that alcohol use does not uniformly lead to sexual risk behavior [20–22]. Even within the same individual, drinking episodes vary: some involve hazardous consumption without sexual risk, some involve near-miss sexual risk (risk is plausible but not enacted), and others culminate in sexual risk behavior [10,23–25]. These patterns suggest that alcohol-related harm reflects not only levels of consumption, but also the processes unfolding within particular drinking episodes. However, the mechanisms through which drinking episodes diverge into these different outcomes remain poorly understood.

Prior quantitative work has examined psychosocial factors (e.g., depression, loneliness), behavioral factors (e.g., substance co-use), and contextual conditions (e.g., social setting) as predictors, moderators, or co-occurring syndemic conditions associated with alcohol use and sexual risk [26–29], but this evidence provides limited insights into how these factors become organized within specific drinking episodes. For example, heavy drinking and condomless sex may co-occur in some relational contexts yet be understood as normative within ongoing partnerships [30], while co-use with other substances can alter pacing, arousal, and perceived risk within episodes [31]. Thus, less is known about how these factors interact dynamically within events to shape whether alcohol-involved episodes escalate toward sexual risk behavior.

Syndemic theory offers a useful framework for examining how co-occurring psychosocial, behavioral, and contextual conditions interact to shape health outcomes [32,33]. Much syndemic research has examined these conditions at the individual or population level, showing how clustered adversities can produce risk beyond single-factor explanations [32,33]. Extending this perspective to the event level allows us to examine how these conditions become activated and organized within discrete drinking episodes. Building on event-level and daily diary research showing that alcohol-involved episodes vary in whether they culminate in SRB [20–25], we use the term *escalation pathway* to describe the sequence through which alcohol-involved events intensify toward potential harm. Prior research suggests that alcohol-related risk may vary according to drinking pace, affective state, social or sexual context, and perceived ability to change course [26,34–36]. We use *interruption of escalation* to describe moments when individual actions, partner or peer responses, protective behavioral strategies, or contextual changes alter, delay, or prevent progression toward SRB [37–40]. Although prior research has examined event-level associations between alcohol use and sexual risk, less is known about how escalation and interruption unfold within specific drinking episodes.

Accordingly, this study examined how alcohol-involved drinking episodes diverged into three outcomes: hazardous alcohol consumption (HAC) only, HAC with near-miss sexual risk, and HAC with sexual risk behavior (SRB). Using a qualitative event-level approach guided by modified grounded theory, we identified within-event escalation pathways and examined how interruption of escalation shaped whether episodes led to HAC only, HAC with near-miss sexual risk, or HAC with SRB.

## Methods

### Study design and approach

We conducted a theory-informed, event-level qualitative analysis of in-depth interview transcripts using a modified grounded theory approach. This approach enabled inductive identification of within-event processes while allowing sensitizing concepts (e.g., syndemic theory) to inform interpretation without prespecifying mechanisms [41–44]. Syndemic theory guided our focus on how biopsychosocial conditions and contextual factors interact within drinking episodes to shape behavior.

Discrete drinking events were the primary unit of analysis. A drinking event was defined as a continuous episode beginning with the initiation of alcohol use and ending when the event terminated (e.g., sleep or next-day resolution). Events were segmented from transcripts as described below.

### Ethical consideration

This study was approved by the University of California, San Francisco Institutional Review Board (IRB protocol #: 23-39758). All participants provided written informed consent before completing the interview. Interviews were audio-recorded with participant permission, transcribed, and de-identified before analysis. Participant quotations were reported using study identifiers to protect confidentiality.

### Data source, sampling, and interviews

Participants were recruited from an ongoing ecological momentary assessment (EMA) parent study examining antecedents and contexts of heavy episodic drinking among adults in the San Francisco Bay Area (SFBA). The parent study administered daily EMA surveys to capture recent alcohol use, contextual factors, and drinking-related experiences. For the present qualitative study, EMA reports of recent drinking episodes provided temporal anchors to support recall during in-depth interviews.

We purposively sampled a subset of EMA participants to capture variation in alcohol-involved episodes, including variation in demographics, co-occurring substance use, and the presence or absence of sexual risk during recent episodes. Eligibility criteria included age ≥18 years, residence in the SFBA, and recent heavy alcohol use. Participants were invited via email, informed about the study and interviewers (e.g., their roles and affiliations), and had no prior relationship with the interviewers. Of 31 invited participants, 22 completed interviews; seven did not respond, and two declined participation. Reasons for non-participation were not systematically tracked.

One-time interviews were conducted via Zoom between June 6 and October 31, 2025, with only the participant and interviewer present. Interviews lasted approximately 30–50 minutes and elicited narratives of participants’ three most recent heavy drinking episodes. Semi-structured prompts asked participants to describe the circumstances before, during, and after each episode, including drinking context, substances used, drinking pace, social and physical setting, relationship or sexual context, perceived risks, and how the episode ended. Participants were also invited to describe behaviors perceived as risky within the event, such as blacking out, substance co-use, condomless sex, or other situations in which alcohol use affected decision-making (S1 Appendix). All participants met a screening threshold of Alcohol Use Disorders Identification Test (AUDIT) score ≥16 [45], indicating moderate to high alcohol-related risk and used in this study to define hazardous alcohol consumption (HAC).

Interviews were conducted by the first author and a trained doctoral research assistant, audio-recorded, transcribed verbatim, and checked for accuracy. The first author had formal training in qualitative methods, including grounded theory. The research assistant received training in qualitative interviewing and conducted interviews under the first author’s supervision. The first author’s perspectives were informed by training and research experience in substance use and syndemic theory; reflexive memos documented assumptions, analytic decisions, and potential biases throughout data collection and analysis. Field notes captured interview context and analytic impressions.

### Event segmentation and data construction

Transcripts were segmented into discrete events beginning with the initiation of alcohol use and ending when the episode terminated. Changes in venue, activity, or interactional context occurring within a continuous drinking episode were treated as part of the same event. A subset of transcripts (25%) was independently reviewed by the second author to confirm event boundaries and event classification; disagreements were resolved through consensus.

Each event was assigned a unique identifier and summarized in a standardized template capturing context, substances used, relational dynamics, and outcomes. The analytic dataset comprised 64 events across 22 interviews (S2 Table).

### Identification of within-event interaction

We distinguished between biopsychosocial conditions (e.g., depression, impulsivity, substance use) and contextual factors (e.g., social environment, relationship dynamics, alcohol availability). Within-event interaction was defined as present when: (1) at least one condition was described as shaping the episode; and (2) another condition or contextual factor interacted with it to influence behavior within the same event. For example, participants described interactions between co-occurring conditions (e.g., depression and cocaine use) or between conditions and contextual factors (e.g., depression and homelessness) that shaped behavior within events.

Classification required evidence of behavioral linkage (e.g., initiating drinking to cope, continuing use to maintain belonging). Co-occurrence without such linkage was not considered sufficient. Events without evidence of interaction were set aside as negative cases for comparison.

This approach reflects an event-level application of syndemic theory, which emphasizes interactions among co-occurring conditions in shaping health outcomes [32,33,46]. To reduce circularity, escalation outcomes (e.g., blackout or sexual risk behavior) were not used as co-occurring conditions and were not used to determine whether events met criteria for within-event interaction. Borderline cases were discussed as a team and documented in analytic memos.

### Analytic strategy and coding procedures

Analysis proceeded using constant comparison, in which events, codes, and emerging categories were iteratively compared within and across cases [41]. We first conducted open coding to characterize events and identify interacting conditions, contextual factors, and initial escalation processes. We then identified recurring escalation pathways and examined how these pathways related to outcomes and interruption within events.

To strengthen analytic rigor and clarify how escalation unfolded within events, we examined how participants described relationships among interacting conditions, escalation processes, and outcomes. Specifically, we assessed whether interactions occurred prior to escalation and whether participants described certain factors as necessary for escalation. We also considered whether alternative factors could plausibly have played a similar role, drawing on the principles of process-oriented analytic approaches [47,48].

### Stage 1: Open coding and event characterization (N=64 events)

The first author conducted open coding at the event level using ATLAS.ti (v25). Codes captured drinking context, substance use, relational dynamics, and immediate outcomes. Events were categorized by outcome as HAC only, HAC with near-miss sexual risk, or HAC with SRB.

Because all participants met the AUDIT threshold for HAC, events were categorized by sexual risk outcomes as: HAC-only episodes, HAC with near-miss sexual risk, or HAC with SRB. HAC-only episodes referred to alcohol-involved episodes among participants with HAC in which no sexual risk was reported. SRB was defined as participant-identified risky sexual activity involving condomless sex with a new or non-monogamous partner, sex with a stranger, or sexual activity occurring under substantial intoxication that could impair consent or protection decisions. Near-miss events were defined as episodes in which such risk was plausible (e.g., opportunity present, intention expressed, or condom negotiation) but did not occur.

Open coding also identified interacting conditions, contextual factors, and instances of interruption (behavioral or contextual regulation within an event that altered escalation).

### Stage 2: Focused interactional analysis and case selection (N=35 events)

Of the 64 events, 35 met criteria for within-event interaction (as defined above) and were included in focused analysis. Events lacking such linkage were excluded from focused analysis but retained as negative cases to refine analytic distinctions. These excluded events typically involved simpler trajectories (e.g., steady drinking without escalation or contextual amplification). Selection was based on analytic criteria rather than outcomes.

Within these 35 events, we examined how interacting conditions shaped escalation and how events unfolded over time. Analysis focused on characterizing escalation within events without prespecifying pathways and outcomes. Additional analytic constructs were developed in subsequent stages through constant comparison.

### Stage 3: Theoretical coding and typology development (N=35)

We synthesized findings into a typology of escalation pathways and mechanisms through iterative comparison. When multiple mechanisms occurred within an event, we distinguished between primary and secondary escalation processes. The primary pathway was defined as the dominant process most proximal to the escalation turning point and most emphasized by the participant’s account of how the event intensified. A secondary pathway or process was defined as a co-occurring process that contributed to escalation but did not serve as the primary driver. Final pathway labels and their roles were developed through iterative comparison and are presented in the Results. Theoretical sufficiency for primary pathways was reached by event 29, with subsequent events confirming these patterns, and no new pathways were identified. Final analysis examined how interacting conditions, escalation pathways, and interruption jointly shaped outcomes.

#### Rigor and Trustworthiness

Study rigor was enhanced through iterative coding and memoing to document analytic decisions, category development, and boundary cases. An audit trail captured event classifications and typology refinement, and event segmentation and classification were independently reviewed for a subset of transcripts, with discrepancies resolved by consensus.

We conducted regular peer debriefings with co-authors to review interpretations. Negative-case comparison using events without evidence of interaction helped refine analytic distinctions and evaluate alternative explanations. Reflexivity was maintained through memos documenting assumptions and interpretive shifts.

This study is reported in accordance with the Consolidated Criteria for Reporting Qualitative Research (COREQ) checklist (S3 Checklist) [49].

### Key analytic constructs

During analysis, we inductively developed analytic constructs through constant comparison to describe how risk unfolded within alcohol-involved events. These constructs were not prespecified; they emerged across analytic stages and were used to organize patterns presented in the Results and depicted in Fig 1. Briefly, *escalation pathways* described recurring processes through which risk increased within events; *escalation constraint point* described the point at which cognitive, relational, or situational constraints reduced participants’ capacity to modify behavior; and *interruption* described behavioral or contextual actions that altered or attempted to alter escalation. The specific escalation pathway types identified through analysis are defined and illustrated in the Results.

**Fig 1.**
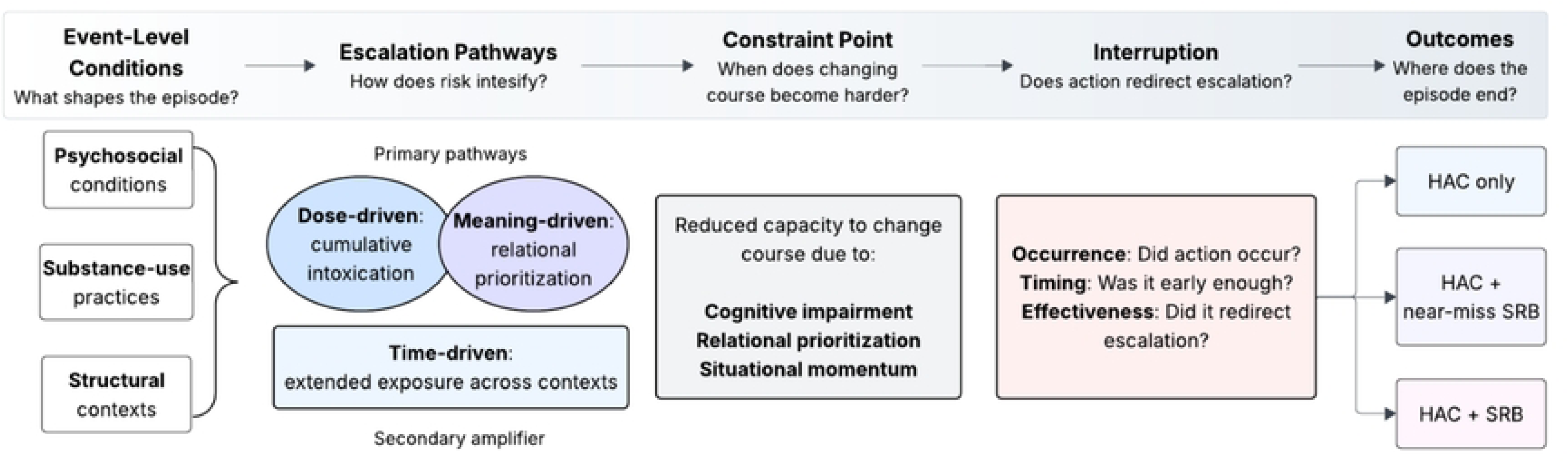
Event-level escalation pathways, constraint, interruption, and outcomes. Psychosocial conditions, substance-use practices, and structural contexts interact within drinking episodes to shape escalation through two primary pathways: dose-driven escalation and meaning-driven escalation. Time-driven continuation functions as a secondary process that extends exposure across contexts and amplifies escalation. As escalation progresses, behavior may reach an escalation constraint point, reflecting cognitive, relational, or situational processes that reduce the capacity to modify behavior. Interruption refers to behavioral or contextual actions that alter or attempt to alter escalation and is characterized by occurrence, timing, and effectiveness. Outcomes reflect whether escalation remains limited to hazardous alcohol consumption, is interrupted prior to sexual risk, or progresses to sexual risk behavior. **Abbreviations:** HAC, hazardous alcohol consumption; SRB, sexual risk behavior.

## Results

### Sample and event distribution

Findings were grounded in participants’ event narratives and illustrated with verbatim quotes. Of the 64 events identified, 35 met criteria for within-event interaction and were included in the focused analysis. The focused analysis examined how co-occurring conditions shaped escalation within these interaction-rich events; events without such interaction were retained as negative cases. Through iterative analysis, we identified recurring escalation pathways and related processes.

The 35 events were reported by 22 participants who were predominantly cisgender men (54.5%), aged 21–49 years (mean 31.7), and primarily identified as heterosexual (86.4%), with racial and ethnic diversity (Table 1). Most participants held at least a bachelor’s degree (68.2%), were employed (72.7%), and over half reported annual incomes ≥ $60,000 (59.1%). All participants reported HIV-negative status.

**Table 1:**
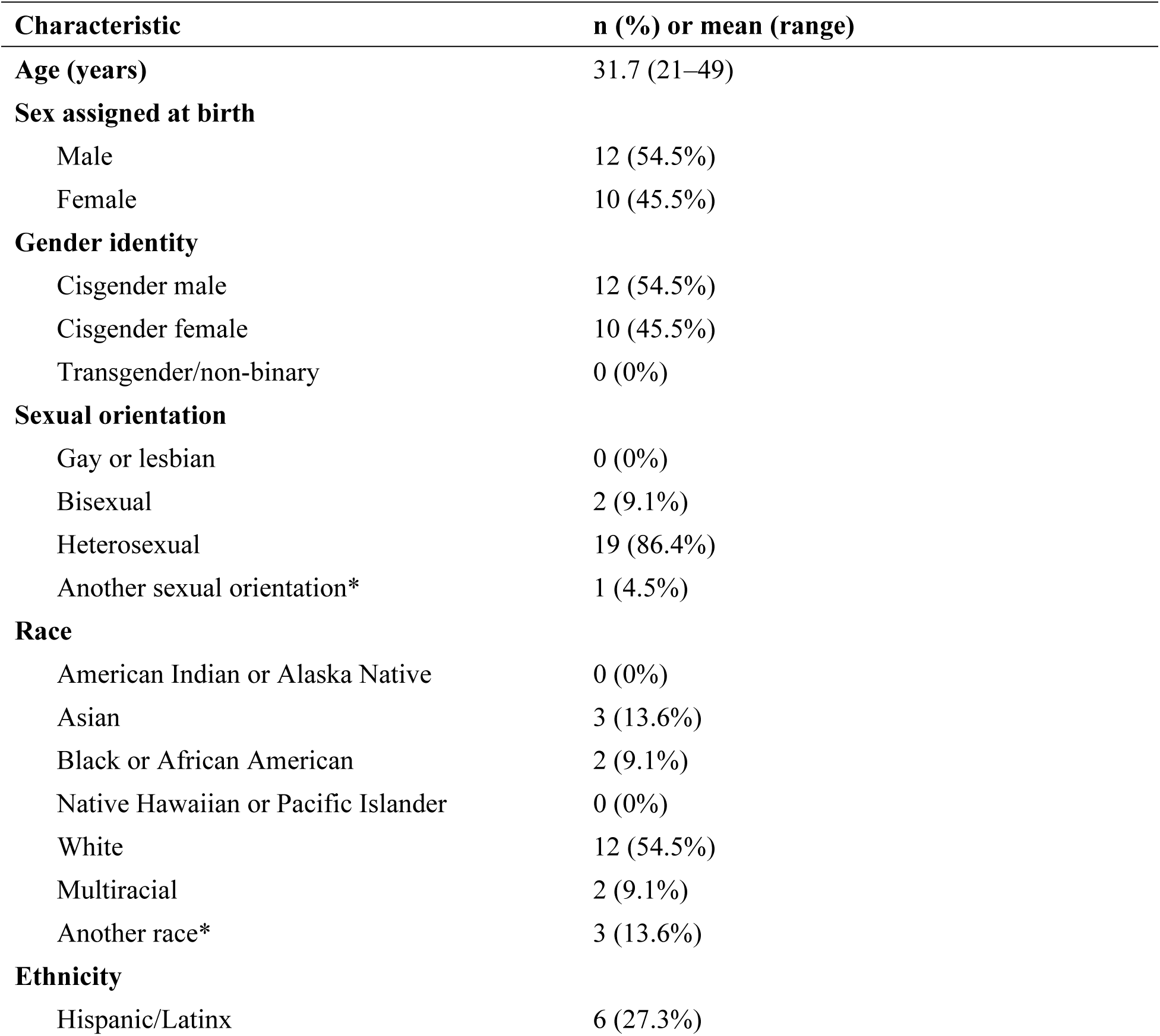
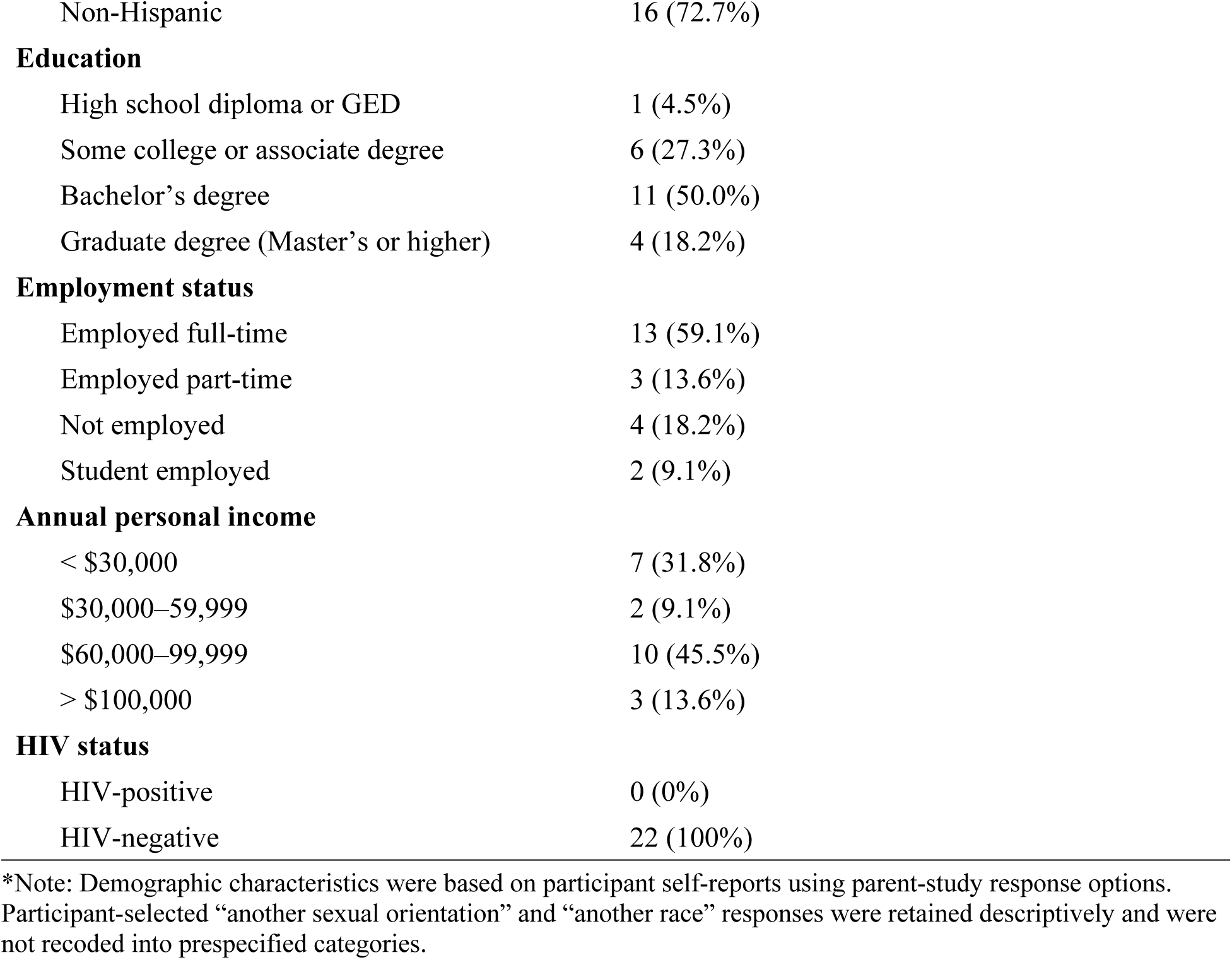
Participant Demographics (N=22)

Of the 35 events included in the focused analysis, 15 (42.9%) resulted in HAC only, six (17.1%) involved HAC with near-miss sexual risk, and 14 (40.0%) involved HAC with SRB.

### Interacting conditions shaping escalation initiation

Escalation initiation involved interacting psychosocial, substance-use, and structural conditions that shaped how participants entered, sustained, and moved through drinking episodes. These conditions did not operate as isolated risk factors. Rather, they became linked within events: psychosocial distress or relational needs shaped motivations for participation, substance-use practices shaped pacing and cumulative exposure, and structural contexts sustained participation across time and settings. These linked conditions provided the event-level foundation for subsequent escalation: cumulative exposure more often organized dose-driven pathways, relational needs often organized meaning-driven pathways, and sustained participation across time and settings, amplifying both through time-driven continuation.

Psychosocial conditions, including distress, relational disruption, and loneliness, shaped motivations for entering and sustaining drinking contexts. Participants described drinking to manage stress (“to cope with those feelings” [8221-1; 8231-1; 8259-1; 8270-3]), numb distress (“dull the senses [and] pain” [8283-1]; “detach [and] numb [their feelings]” [8250-1]), or facilitate social engagement (“lubricate myself for social events” [8262-3]). Relational loss and loneliness further motivated efforts to “connect [with others]” (8231-2; 8270-2; 8283-1), seek “attention [and] validation” (8221-2; 8278-2), or drink “for the sake of not being alone” (8225-1).

Substance-use practices shaped cumulative exposure through pacing, sequencing, and co-use. Participants described sustained drinking to maintain intoxication (e.g., avoid “losing my buzz.” [8250-3]). Co-use often followed initial intoxication (“once we were tipsy or drunk, then the cocaine comes out” [8210-1]) or was used to offset fatigue (“If I feel tired…get some more cocaine” [8210-1]; “cocaine makes me more alert” [8274-2]). These practices were framed in relational (“people just love giving me drugs” [8225-2]), emotional (“it felt nice to smoke while drinking” [8221-2]), and behavioral terms (“I’ll have a gummy so I’m not drinking as much” [8250-2]).

Structural contexts sustained participation across time and settings, embedding drinking within extended social flow. Participants described multi-stage sequences (“friend’s house…then the bar…then a Bloody Mary the next day” [8262-3]) and prolonged episodes (“midnight until nine in the morning” [8259-3]). Conformity pressures (“fit in with the team” [8266-2]; “prove I could drink” [8283-3]), situational cues (“If you put a shot in front of me…I’ll be like, ‘Okay, screw it!’” [8221-1]), and internalized norms (“In order to have fun, I need to drink” [8210-2]; “I don’t like not having something in my hands” [8219-2]) further reinforced ongoing consumption.

Together, these interacting conditions linked participants’ motivations, substance-use practices, and drinking contexts, forming the event-level foundation for the escalation pathways described below.

### Escalation pathways

Through iterative comparison across events, we identified two primary escalation pathways: dose-driven and meaning-driven. A third process, time-driven continuation, functioned as a secondary process that extended exposure across contexts and amplified dose-driven or meaning-driven escalation (Tables 2 and 3). Pathways could co-occur within events; however, each event was assigned one primary pathway based on the process most proximal to the escalation turning point and most emphasized in the participant’s account.

**Table 2:** Primary escalation pathways and interruption patterns across drinking events (*N*=35)

| Escalation<br>Pathway | Interruption before<br>constraint and altered<br>escalation<br>*n* (%) | Interruption after constraint<br>and/or did not alter<br>escalation<br>*n* (%) | No Interruption<br>*n* (%) | Total<br>*n* (%) |
| --- | --- | --- | --- | --- |
| Dose-driven | 3 (8.6%) | 2 (5.7%) | 16 (45.7%) | 21 (60.0%) |
| Meaning-driven | 3 (8.6%) | 3 (8.6%) | 8 (22.9%) | 14 (40.0%) |
| Total | 6 (17.1%) | 5 (14.3%) | 24 (68.6%) | 35 (100.0%) |

**Note.** Percentages are based on 35 focused events. Primary escalation pathway refers to the dominant process most proximal to the escalation turning point within an event. Interruption was characterized into three dimensions: occurrence (whether the action occurred), timing (relative to the escalation constraint point), and effectiveness (whether the action altered escalation). Time-driven continuation did not occur as a primary pathway; it functioned as a secondary process that extended exposure across contexts and amplified dose-driven and meaning-driven escalation. Secondary pathways are presented in the S4 Table.

**Table 3:** Event-level escalation pathways and mechanisms in drinking events (*N*=35)

| Escalation Pathway | Description | Mechanisms | Constraint Point | Narrative Indicators | Potential Interruption Point | Exemplar Quotes |
| --- | --- | --- | --- | --- | --- | --- |
| Dose-driven | Escalation driven by cumulative alcohol and/or substance exposure, resulting in progressive impairment awareness and self-regulation | Threshold drift: intended limits exceeded through rapid stacking or compressed dosing.<br>Exposure prolongation: alcohol or substance use continued after limits exceeded or through sustained co-use. | Impairment: reduced awareness and executive control | Exceeding intended limits without awareness; recognizing risk only after intoxication; reduced follow-through; blackout or near-blackout; continued use despite intoxication | Before substantial impairment, when participants still had capacity to slow, stop, leave, or seek support. | “I was definitely thinking, ‘This is probably not the best idea,’ but didn’t follow through.” [8286-1] |
| Meaning-driven | Escalation driven by prioritizing connection, intimacy, validation, or belonging despite awareness of risk | Relational framing: drinking or sexual behavior was organized around validation seeking, intimacy preservation, belonging display. | Relational prioritization: relational goals become prioritized over risk reduction or protective decision-making | Awareness or risk with continued participation; avoiding disruption of connection; condomless sex framed as intimacy; drinking framed as facilitating connection | Before relational goals became prioritized over risk reduction, when boundaries, condom negotiation, or disengagement remained more feasible. | “I knew we should use a condom, but I didn’t want to ruin the moment.” [8274-2] |
| Time-driven | Secondary process in which continued participation across contexts extended exposure and amplified dose-driven and meaning-driven escalation | Momentum locking: venue transitions, chaining events, or sequential extension prolonged participation and reduced opportunities to disengage. | Situational commitment: embedded continuation across contexts | Sequential context changes; repeated postponement of leaving; “one more” continuation; escalation without explicit intent; feeling committed or “too far in” | Before situational commitment formed, when participants could still exit, avoid the next setting, or interrupt the sequence. | “I kept thinking I’d go home after the next stop, but we just kept going.” [8262-1] |

**Note:** Escalation pathways and mechanisms were derived through constant comparison across 35 events. Dose-driven and meaning-driven escalation were primary pathways when they served as the dominant mechanism most proximal to the escalation turning point. Time-driven continuation functioned as a secondary process that extended exposure across time or settings and amplified primary pathways but did not function as the dominant pathway in this dataset. The potential interruption point indicates when an interruption was most likely to alter escalation before the relevant constraint point was reached.

Table 2 summarizes how primary escalation pathways intersect with interruption patterns. Dose-driven escalation was the primary pathway in 21 events (60.0%); most of these events had no interruption (*n*=16), while fewer involved interruption before the constraint point that altered escalation (*n*=3) or interruption after the constraint point and/or interruption that did not alter escalation (*n*=2). Meaning-driven escalation was the primary pathway in 14 events (40.0%) and showed a similar pattern, with most events having no interruption (*n*=8).

Table 3 summarizes the escalation pathways and mechanisms identified through constant comparison. Dose-driven and meaning-driven escalation emerged as primary pathways, whereas time-driven continuation functioned as a secondary process that extended exposure across contexts and amplified either pathway. The table shows how each pathway was identified in participants’ narratives, the point at which behavior became more difficult to modify, and when interruption appeared most likely to alter the trajectory. These pathways also helped explain divergent outcomes: HAC-only events more often reflected dose-driven escalation, SRB events more often reflected meaning-driven escalation, and near-miss events illustrated how earlier interruption could redirect escalation before sexual risk occurred.

#### Dose-driven pathway

Dose-driven escalation involved cumulative intoxication that progressively impaired awareness and self-regulation. Progression was driven by pharmacological exposure (i.e., alcohol and other substances) rather than relational priorities, with escalation emerging through threshold drift and prolonged use.

Threshold drift occurred when intended limits were exceeded without clear awareness, often due to rapid stacking or compressed dosing. What began as moderate drinking escalated to “10-ish drinks” (8266-2) by “cramming too many drinks into too short a time.” (8274-1) Fatigue sometimes compounded intoxication (“I hadn’t slept…then drank a lot and blacked out” [8219-1]). Participants described reduced inhibition and deliberation (“my inhibition had been lowered” [8270-1]; “I wasn’t thinking through my decisions” [8278-3]).

Exposure prolongation occurred when drinking or co-use continued after limits were exceeded. In this sample, cocaine was the most frequently co-used substance, often routinized with alcohol (“I was doing a lot of cocaine…evolved into cocaine and alcohol abuse” [8225-2]; “I don’t think I’ve ever used cocaine without alcohol” [8274-2]). Combined use amplified disinhibition and fatigue (“the combination of alcohol and cocaine makes me very talkative” [8270-2]; “I was running on fumes.” [8225-1]) and culminated in severe impairment, including blackout (“blacking out [from mixing alcohol and cocaine]” [8259-3]). Escalation in these events reflected progressive impairment.

#### Meaning-driven pathway

Meaning-driven escalation involved prioritizing connection, intimacy, or a sense of belonging despite awareness of risk. In these events, drinking and sexual behavior functioned as means to achieve relational goals rather than being driven by intoxication alone.

Alcohol use intensified as interactions became relationally salient, including efforts to connect (“getting closer [to partner]” [8249-3]; “[using] alcohol to connect” [8231-1]), cope with relational distress (“drinking to cope [after seeing ex-partner]” [8221-1]), seek validation (“male attention” [8278–2]), manage social anxiety (“drink to relax” [8231-1]; “warm myself up” [8262-1]), and avoid loneliness (“for the sake of not being alone” [8225-1]).

Drinking also functioned in belonging and identity displays (“prove that I could hang with [peers]” [8283-3]) and conformity (“going with whatever the rest of the crowd was doing” [8262-2]). Occupational roles (“[clients] started buying me shots” [8221-3]) and structural stressors further reinforced this pathway (“drinking helps me detach from stress…” [8250-1]; “being a single parent and losing my job…did lead me to drink more” [8254-1]).

Even when risk was recognized, participants deprioritized precaution to preserve the relational goals (“I knew we should use a condom, but I didn’t want to ruin the moment” [8274-2]; “I just wanted to feel wanted” [8225–2]). Escalation in these events reflected relational prioritization rather than impaired awareness.

#### Time-driven continuation (secondary process)

Time-driven continuation extended exposure across contexts and reduced opportunities for interruption. Although not a primary pathway, it amplified both dose- and meaning-driven escalation by embedding participation over time.

Small extensions, such as venue changes or “one more stop,” accumulated into sustained exposure. Participants described chained contexts (“bar…then his place…kept drinking” [8239-1]; “one party to another party and then back to someone’s apartment” [8286-1]) and ongoing social flow (“every time my drink got down, she filled it again” [8250-1]; “company party with open bar…once the alcohol starts flowing…I was out of control” [8249-3]).

Exit was repeatedly postponed (“I kept thinking I’d go home after the next stop, but we just kept going” [8262-1]) until continuation became embedded (“already too far into the night to leave” [8278-1]). In these cases, situational commitment constrained interruption even when cognitive awareness remained intact.

### Escalation constraint point

Across pathways, a recurring process marker, termed as the escalation constraint point, emerged as the stage at which participants’ capacity to modify behavior became reduced due to accumulating constraints. This point reflects the convergence of cognitive (impairment), relational (prioritization of connection), and situational (momentum across contexts) processes that limit behavioral flexibility and reduce the feasibility of interruption.

In participants’ narratives, constraint points appeared as moments when their ability to change course narrowed. These moments involved cognitive constraints (e.g., impaired decision-making), relational constraints (e.g., reduced ability to assert preferences or disengage), and situational constraints (e.g., inability to leave or disengage from ongoing situations). Participants described loss of cognitive control (“I did black out…I don’t remember all of it [8259-1]), reduced agency (“I just went with [the unprompted sex]” [8225-2]), and sustained situational momentum (“[The drinks] just keep coming” [8249-3]).

Although all three forms of constraint could co-occur within events, their relative prominence differed across pathways. In dose-driven pathways, constraint was primarily experienced as cognitive impairment that reduced awareness and behavioral control. In meaning-driven pathways, constraint reflected relational prioritization over risk reduction. In time-driven continuation, constraints emerged through sustained participation across contexts that limited opportunities for disengagement. Across pathways, these processes often converged, and participants typically recognized risk only after constraint had formed, limiting the feasibility of behavioral change.

### Interruption

We then examined how interruption operated within events, focusing on when actions occurred relative to the escalation constraint point and whether they altered escalation. Interruption varied along three dimensions: occurrence (whether action occurred), timing (when it occurred relative to the escalation constraint point), and effectiveness (whether it altered escalation).

Across events, recognition of risk typically occurred after the escalation constraint point had formed, limiting the feasibility of effective interruption. In dose-driven pathways, recognition followed impairment (“I was definitely thinking [not wearing a condom] is probably not the best idea but didn’t follow through on the impulse” [8286-1]). In meaning-driven pathways, risk was acknowledged but deprioritized in favor of relational goals (“I knew we should use a condom, but I didn’t want to ruin the moment” [8274-2]). In time-driven continuation, awareness emerged after participants were already engaged in ongoing social situations or sequences of events (“we just kept going” [8262-1]). Recognition alone rarely resulted in interruption once escalation had progressed.

When an interruption occurred before the escalation constraint point and altered escalation, trajectories were more likely to remain contained. Participants described leaving the setting (“I realized I was really drunk, so I just took an Uber home” [8221-3]) or asserting boundaries (“I told him to stop and that we should just go to bed” [8210-1]), substituting substances to limit drinking (“I’ll have a gummy so that I’m not drinking as much” [8250-2]), or disengaging from interaction (“he was being a little persistent…and I kind of put him in his place at one point” [8221-2]). These actions redirected the trajectory of escalation.

In contrast, when an interruption occurred after the constraint point and/or did not alter escalation, events were more likely to progress. Participants described monitoring without effective regulation (“I try to watch everything I do” [8254-2]) or partial boundary management (“I don’t let him ejaculate inside me” [8239-1]) that reduced but did not prevent progression. In some cases, these efforts were overridden by emotional or relational dynamics, culminating in risky behavior (“having rough sex [without a condom]” [8254-2]).

Other events were characterized by the absence of interruption, with escalation continuing despite ongoing exposure and, in some cases, awareness of risk (“the mixture [of alcohol and cocaine] is where I began to black out” [8259-3]; “five to ten more…at a certain point, I don’t recall even drinking those drinks” [8262-1]).

Across events with similar levels of intoxication, outcomes diverged according to when the interruption occurred and whether it altered escalation. Early and effective interruption, occurring before the constraint point, was associated with trajectories that remained contained. In contrast, an interruption that was absent, occurred after the constraint point, or did not alter escalation was associated with progression to HAC or SRB.

### Outcomes across pathways

Outcomes differed across escalation pathways and interruption patterns. HAC-only events more often followed dose-driven pathways, whereas events involving SRB more often followed meaning-driven pathways. Near-miss events occurred across both pathways and were characterized by an interruption occurring before the escalation constraint point.

These patterns indicate that similar levels of alcohol use reported by participants may lead to different outcomes depending on how escalation unfolds and whether interruption occurs before constraint formation. Near-miss events, although limited in number (n = 6), were analytically informative in illustrating how participants recognized risk and modified behavior before escalation progressed to SRB. Across events, divergence in outcomes reflected the sequence linking interacting conditions, escalation pathways, constraint formation, and interruption, rather than differences in the amount of alcohol consumed alone.

## Discussion

This study examined how alcohol-involved drinking episodes diverged into three outcomes: HAC only, HAC with near-miss sexual risk, and HAC with SRB. The central finding is that alcohol exposure alone did not account for these divergent outcomes; rather, outcomes appeared to depend on how escalation unfolded within events and whether interruption occurred before an escalation constraint point. HAC-only events more often reflected escalation centered on cumulative intoxication rather than sexual-risk dynamics, near-miss events illustrated moments when participants recognized risk and altered the trajectory before SRB occurred, and SRB events more often reflected escalation after relational prioritization or situational commitment had constrained participants’ ability to change course. This pattern helps explain why similarly described intoxication narratives can yield different sexual-risk outcomes and aligns with event-level and daily diary research showing that alcohol-related sexual behavior varies by drinking context, day-level alcohol use, and relational factors [26,28,30].

Escalation unfolded as a temporal sequence linking interacting event-level conditions, pathway-specific processes, an escalation constraint point, and interruption. Within this sequence, dose-driven and meaning-driven pathways organized escalation, often amplified by time-driven continuation. Thus, outcomes depended not only on whether interruption occurred but also on when it occurred and whether it altered the trajectory.

These findings extend syndemic theory by showing how interacting psychosocial, substance-use, and contextual conditions can operate within discrete drinking episodes rather than functioning only as background vulnerabilities [32,33,46]. In this study, these conditions shaped event-level processes through which episodes unfolded, including motivations for participation, pacing and continuation of substance use, relational priorities, and opportunities for disengagement. Structural factors, including drinking environments, social norms, and access to substances, shaped entry into escalation pathways and sustained participation across contexts, even when they did not determine pathway type. This event-level perspective helps clarify how interacting syndemic conditions may translate into different episode trajectories, including HAC-only events, near-miss sexual risk, and SRB.

The escalation typology suggests that hazardous drinking episodes are organized by proximal event-level processes rather than by intoxication magnitude alone. Dose-driven and meaning-driven pathways differed in the processes that moved episodes toward risk and in the constraint points that made interruption more or less feasible. This distinction suggests that sexual risk in drinking episodes may arise not only from cumulative pharmacological exposure but also from relational processes through which drinking, intimacy, validation, or belonging become linked within the event [11,28,30].

Time-driven continuation functioned as a secondary process that embedded participation across sequential contexts and further reduced opportunities for interruption. Although not a primary pathway in this dataset, its recurrent co-occurrence suggests that continued participation across contexts amplified dose- and meaning-driven pathways by extending exposure duration and delaying disengagement. This finding aligns with prior event-level studies demonstrating that drinking contexts and venue transitions contribute to acute alcohol-related harms independent of individual risk characteristics [26,29].

Across events, escalation constraint points marked moments when participants’ ability to act on intentions to reduce risk narrowed substantially. These points reflected the convergence of cognitive impairment, relational prioritization, and situational momentum. Participants often recognized risk but did not prevent progression because recognition occurred after the episode had already become difficult to redirect. This temporal misalignment helps explain why heavy drinking does not uniformly co-occur with sexual risk within individuals or across episodes [10,20,50]. It is also consistent with alcohol myopia and acute alcohol effects models, which suggest that intoxication can narrow attention, weaken behavioral regulation, and alter sexual decision-making [11,51].

Another key insight from this study is that interruption, defined by its occurrence, timing, and effectiveness, functions as a proximal mechanism linking escalation pathways to outcomes. Participants frequently recognized risk but acted only after impairment, relational prioritization, or situational commitment had formed, when interruption was less feasible. In contrast, actions taken earlier, such as leaving settings, asserting boundaries, or limiting intake, were more likely to alter escalation trajectories. These findings suggest that alcohol-related harm may depend less on knowledge or intention than on whether action occurs before escalation constraints accumulate within episodes [10,11,24]. This highlights the potential importance of prevention strategies that support earlier recognition, boundary-setting, pacing, exit planning, and peer or partner intervention before behavior becomes difficult to redirect.

### Implications

Many alcohol-related sexual-risk interventions focus on the transition from drinking to sexual behavior; in contrast, escalation in this study often began earlier within drinking episodes [17,24]. Prevention efforts may be most useful when they support action before the escalation constraint point, when episodes may still be redirected toward HAC-only or near-miss trajectories rather than progressing to SRB. Examples include supporting pacing before impairment, reinforcing relational boundaries before relational prioritization, or enabling exit before individuals become embedded in ongoing situations or sequences of events.

These findings highlight intervention strategies that align with harm-reduction approaches, including pre-event harm reduction planning, early-episode mobile supports, and structural supports that make leaving easier, such as transportation access or peer monitoring. Such approaches shift emphasis from modifying individual risk traits to shaping situational conditions that enable timely action within episodes [29].

### Limitations

The sample comprised primarily heterosexual cisgender adults in the San Francisco Bay Area with relatively high socioeconomic status, limiting generalizability to populations with different structural conditions or risk profiles. Escalation pathways may operate differently in populations facing structural marginalization, where stigma, economic instability, and constrained access to resources may shape relational dynamics and opportunities for interruption. These dynamics were not directly examined and warrant further investigation.

Event narratives relied on retrospective recall, although participation in ecological momentary assessment likely improved temporal anchoring. The analytic sample was restricted to events demonstrating within-event interaction; thus, the identified pathways reflect processes under conditions of interaction and may not characterize all alcohol-involved episodes. Counts within pathway–outcome combinations were small and should be interpreted descriptively. The small number of near-miss events limits the strength of inferences for this subgroup; findings should be interpreted as exploratory.

## Conclusion

Drinking episodes diverge into hazardous consumption with or without sexual risk through interacting mechanisms that shape how escalation unfolds within events. Dose-driven and meaning-driven pathways represent distinct processes through which event-level conditions shape behavior, culminating in escalation constraint points that limit the feasibility of interruption. Outcomes depend on whether the interruption occurs and when it occurs relative to the escalation constraint point. An event-level perspective, therefore, identifies early stages of escalation, prior to impairment, relational prioritization, or sustained participation across contexts, as critical opportunities for harm-reduction intervention.

## Data Availability

The data underlying this study consist of qualitative interview transcripts. Relevant de-identified excerpts supporting the findings are included within the manuscript. Full interview transcripts cannot be made publicly available because they contain sensitive information about participants’ alcohol and substance use, sexual behavior, and personal social contexts that could compromise participant confidentiality. Public sharing of full transcripts was not included in the informed consent process and is restricted by the University of California, San Francisco Institutional Review Board. Additional de-identified excerpts may be made available upon reasonable request to the corresponding author, subject to UCSF IRB approval and applicable data-use requirements.

## Acknowledgments

We thank Claire Draucker (Indiana University, School of Nursing) for methodological guidance in designing the modified grounded theory study and Michael Deynu (University of California, San Francisco, School of Nursing) for assistance with participant interviews.

## Supporting information

**S1 Appendix. Qualitative interview guide.** Semi-structured interview guide used to elicit participant narratives of recent heavy drinking and/or sexual-risk events, including prompts about event context, sequence, co-occurring challenges, substance co-use, coping, and perceived support needs.

**S2 Table. Event-level analytic summary of segmented drinking episodes.** De-identified summary of the 64 drinking events segmented from participant interviews, including manuscript outcome category, substance co-use, inclusion in the focused interactional analysis, brief event summary, and analytic classification.

**S3 Checklist. Consolidated Criteria for Reporting Qualitative Research checklist**. Completed COREQ 32-item checklist documenting reporting of the research team and reflexivity, study design, data collection, analysis, and findings.

**S4 Table. Escalation pathway roles and interruption patterns across focused drinking events.** Distribution of primary and secondary escalation pathway roles and interruption patterns across the 35 events included in the focused interactional analysis.

## Notes

### Competing Interest Statement

The authors have declared no competing interest.

### Funding Statement

Yes

### Author Declarations

The parent study and qualitative interview procedures were approved by the University of California, San Francisco Institutional Review Board (UCSF IRB IRB number: 23-39758)

